# Test-retest reliability and concurrent validity of the Adapted Short QUestionnaire to ASsess Health-enhancing physical activity (Adapted-SQUASH) in adults with disabilities

**DOI:** 10.1101/2020.09.09.20190371

**Authors:** Bregje L. Seves, Femke Hoekstra, Jorrit W.A. Schoenmakers, Pim Brandenbarg, Trynke Hoekstra, Florentina J. Hettinga, Rienk Dekker, Lucas H.V. van der Woude, Cees P. van der Schans

**Affiliations:** University of Groningen, University Medical Center Groningen, Center for Human Movement Sciences, Groningen, The Netherlands.; University of Groningen, University Medical Center Groningen, Department of Rehabilitation Medicine, Groningen, The Netherlands.; University of British Columbia Okanagan, School of Health and Exercise Sciences, Kelowna, British Columbia, Canada.; University of Groningen, University Medical Center Groningen, Department of Orthopedics, Groningen, The Netherlands.; Vrije Universiteit Amsterdam, Department of Health Sciences and Amsterdam Public Health research institute, Amsterdam, The Netherlands.; Northumbria University, Department of Sport, Exercise and Rehabilitation, Newcastle, United Kingdom.; Hanze University of Applied Sciences, Research and Innovation Group in Health Care and Nursing, Groningen, The Netherlands.; Peter Harrison Centre for Disability Sport, School of Sport, Exercise & Health Sciences, Loughborough University, UK.

**Keywords:** Physical activity assessment, accelerometer, chronic disease, rehabilitation, health promotion

## Abstract

The current study determined the test-retest reliability and concurrent validity of the Adapted Short QUestionnaire to ASsess Health-enhancing physical activity (Adapted-SQUASH) in adults with disabilities. Before filling in the Adapted-SQUASH twice with a recall period of two weeks, participants wore the Actiheart activity monitor up to one week. For the test-retest reliability (N = 68), Intraclass correlation coefficients (ICCs) were 0.67 (p < 0.001) for the total activity score (min x intensity/week) and 0.76 (p< 0.001) for the total minutes of activity (min/week). For the concurrent validity (N = 58), the Spearman correlation coefficient was 0.40 (p = 0.002) between the total activity score of the first administration of the Adapted-SQUASH and activity energy expenditure from the Actiheart (kcals kg^−1^ min^−1^). The ICC was 0.22 (p = 0.027) between the total minutes of activity assessed with the first administration of the Adapted-SQUASH and Actiheart. The Adapted-SQUASH is an acceptable measure to assess self-reported physical activity in large populations of adults with disabilities, but is not applicable at the individual level due to wide limits of agreement. Self-reported physical activity assessed with the Adapted-SQUASH does not accurately represent physical activity assessed with the Actiheart in adults with disabilities, as indicated with a systematic bias between both instruments in the Bland-Altman analysis.

## Introduction

Measuring patients’ physical activity behaviour is important for evaluating effectivity of physical activity promotion interventions and, ideally, individually tailoring rehabilitation programmes among adults suffering from a physical disability and/or chronic disease that impairs mobility (further: adults with disabilities) [1]. Therefore, an accurate and efficient measurement instrument for assessing (self-reported) physical activity in people with physical disabilities is essential. Although accelerometer-derived physical activity might be preferred, mostly it is not practically feasible, and it is too expensive among large scale populations in interventions and/or cohort studies. Self-reports are frequently used measurement tools to assess physical activity in disabled populations, both in rehabilitation practice and in research [2,3]. Also, questionnaires are easy to fill in [4–6]. However, self-reported physical activity depends on the persons’ recall and mostly is not sensitive for light physical activities at home (e.g. walking from the bedroom to the toilet and from the kitchen to the dining table) or outside (e.g. walking to the mailbox to post a letter) [7].

A self-reported physical activity measure was needed in the multicentre longitudinal cohort study Rehabilitation, Sports and Active lifestyle (ReSpAct) to evaluate physical activity during and after the physical activity stimulation programme Rehabilitation, Sports and Exercise (RSE; Dutch: ‘Revalidatie, Sport en Bewegen’) [8,9]. The RSE programme was successfully implemented in eighteen rehabilitation institutions in the Netherlands [10]. The questionnaire was required to be suitable for the target population: people with a physical disability and/or chronic disease. There are few physical activity questionnaires available specifically developed for adults with disabilities [3]. The Physical Activity Scale for Individuals with Physical Disabilities (PASIPD) [11] was considered for the ReSpAct study, since it is commonly used amongst the target population. To precisely assess the physical activity behaviour before and after a physical activity promotion intervention [6], and to clarify the dose-response relationship between physical activity and the received counselling during the intervention [12], frequency, intensity, duration and type of the activity should be measured. The PASIPD assesses duration and type of physical activities but does not specifically assess the frequency and intensity of physical activities, whereby it was considered not applicable for the ReSpAct cohort. The Short Questionnaire to Assess Health-enhancing physical activity (SQUASH) developed for healthy adults does measure frequency, intensity, duration and type of physical activities [4]. The SQUASH is widely used, for example by governmental agencies to monitor large scale physical activity behaviour among the Dutch population and to monitor whether physical activity guidelines are achieved. Studies on the psychometric properties of the SQUASH have supported the appropriateness of the SQUASH to measure the level of weekly physical activity in a healthy adult population [4], in patients after a total hip arthroplasty [13] and in outpatients with ankylosing spondylitis [14].

When assessing physical activity in adults with disabilities, it needs to be taken into account that this target population may have a different perceived intensity of activities compared to a healthy population [15]. It is expected that adults with a disability experience activities as more intense, because activities often cost (absolutely and relatively) more energy compared to healthy adults [16,17]. Therefore, the ReSpAct research team converted the original SQUASH into a measurement tool (mentioned from here: the Adapted-SQUASH) that was expected to better meet the perceived intensity of activities among people with disabilities compared with the original SQUASH, by using appropriate metabolic equivalent of task (MET) values for this target population. Also, the SQUASH was adapted to better match the activity pattern of wheelchair users by including common physical activity behaviours: wheelchair sports (e.g. wheelchair basketball) and questions concerning wheelchair propulsion and handcycling [9]. The SQUASH has two main outcome measures: the activity score, measuring a combination of intensity and duration of physical activity per week, and total minutes of activity per week (duration).

It is relevant for (rehabilitation) practice and research in (adapted) physical activity to determine the psychometric properties of the Adapted-SQUASH among a sample of adults with disabilities. Apart from test-retest reliability, construct validity is deemed an important asset. The Actiheart (Cambridge Neurotechnology™ UK), an uniaxial activity monitor, was identified by the research team as a suitable criterion measure to compare with the outcomes of the Adapted-SQUASH. Since the Actiheart is a medical device, it is suitable for ambulant people with disabilities. The Actiheart is accurate in measuring physical activity energy expenditure (AEE) in free living conditions, and ideally it combines its measured heart rate and movement sensor information improving the prediction of AEE in daily physical activities [5,18].

The current study aims to determine the test-retest reliability and concurrent validity of the Adapted-SQUASH among adults with disabilities. We focused on the two main outcome measures of the Adapted-SQUASH, the total activity score and the total minutes of activity per week [4], which were derived from the test and retest of the Adapted-SQUASH as well as from the Actiheart activity monitor among a convenience sample of adults with disabilities.

## Methods

### Study population

Participants were recruited through patient activity groups in hospitals, rehabilitation centres, sport clubs and patient associations in the northern and eastern provinces in the Netherlands. Inclusion criteria were being at least eighteen years of age, having a physical disability and/or chronic disease (e.g. stroke, heart failure, Parkinson’s disease) and being able to read and write the Dutch language. Participants were excluded when they were still receiving inpatient or outpatient rehabilitation care, were participating in the ReSpAct study [8,9], were completely wheelchair dependent (because of the use of the Actiheart), or were not able to complete the questionnaires even with help. The data collection took place from November 2014 till June 2016.

### Study procedures

This study consisted of a test-retest reliability study and a validity study. For the test-retest reliability study, the participants filled out the first Adapted-SQUASH twice, with approximately two weeks between the measurement occasions.

For the validity study, the participant was asked to wear an Actiheart activity monitor (Cambridge Neurotechnology™ UK) to objectively measure physical activity levels during the week prior to administration of the first Adapted-SQUASH. Two researchers visited the participants in their free-living home situation twice, to install and attach the Actiheart to the participants’ chest, and to collect the Actiheart after one week. The Actiheart measurement started at 00:00 AM and continued for the next seven consecutive days, both day and night. The participant was instructed to remove the Actiheart during showering, bathing, or swimming. In addition, the participant filled out a diary in which noncompliance to the Actiheart was noted. Measurements were included in the validity study when a minimum registration of the Actiheart of at least four days valid acceleration data (at least 75% activity data registration of 24 hours) for each participant was present [19].

Participants’ general characteristics were obtained by using a questionnaire. Participant’s body weight (kg) and height (m) were measured by researchers by using a personal scale and measuring tape, respectively. The study was approved by the Ethics Committee of the Center of Human Movement Sciences (ECB/2014.06.30_1) at the University of Groningen, University Medical Center Groningen. All participants voluntarily signed an informed consent.

### The Adapted Short QUestionnaire to ASssess Health-enhancing physical activity (Adapted-SQUASH)

The 19-item Adapted-SQUASH (see supplemental file) is a self-reported recall questionnaire to assess physical activity among adults with disabilities based on an average week in the past month as reference period. Equal to the original SQUASH [4], the Adapted-SQUASH is pre-structured in four main domains outlining types and settings of activity: ‘commuting traffic’, ‘activities at work and school’, ‘household activities’ and ‘leisure time activities’ including ‘sports activities’. The frequency in days per week, the duration in average hours and minutes per day and the perceived intensity were asked.

Several adjustments have been made to make the original SQUASH applicable for people with disabilities, as described in the study protocol of the ReSpAct study [9]. First, the items ‘wheelchair riding’ and ‘handcycling’ were added in the domains ‘commuting activities and leisure-time’ and ‘sports activities’. Second, the self-reported intensity of the activity was categorised into ‘light’, ‘moderate’ and ‘vigorous’, instead of ‘slow’, ‘moderate’ and ‘fast’. Third, the syntax to determine the outcome measures of the Adapted-SQUASH includes a large range of Adapted sports (e.g. wheelchair basketball/rugby/tennis) for the item ‘sports activities’. The MET-values in the syntax were updated based on the most recent version of the Ainsworth’ compendium of physical activities [20] and MET-values for wheelchair riding, handcycling and adapted sports were added based on a compendium of energy costs of physical activities for wheelchair dependent individuals [21]. Lastly, in the examples of different sports ‘tennis’ was replaced by ‘(wheelchair) tennis’.

### The total activity score per week (Adapted-SQUASH)

For practical use of the questionnaire all outcome measures of the Adapted-SQUASH were calculated by using a syntax. The total activity score and the total minutes of activity per week are the main outcomes of the Adapted-SQUASH. The total activity score (min x intensity/week) was calculated following the procedure described by Wendel-Vos et al. (2003). First, all the questions in the Adapted-SQUASH were assigned to a MET-value representing the intensity of this task, based on the Ainsworth’ compendium of physical activities [20] and based on a compendium of energy costs of physical activities for wheelchair dependent individuals [21]. Second, an activity score was calculated for each domain by multiplying the total minutes of activity with a self-reported intensity score, which is based on age and MET-values [4]. Lastly, the total activity score was calculated by summing up the activity scores of the four domains. In accordance with the original SQUASH, data were excluded if the total minutes of activity a day exceeded 960 minutes or if values were missing [4].

### The total minutes of activity per week (Adapted-SQUASH)

The total minutes of activity per week (min/week) assessed with the Adapted-SQUASH were calculated by summing up the total minutes of physical activity per week reported in the Adapted-SQUASH. Also, the total minutes of light, moderate and vigorous intensity activities per week (min/week) were calculated, using MET-value cut-off points based on the Dutch physical activity guidelines [22].

### The Actiheart activity monitor

The Actiheart (Cambridge Neurotechnology™ UK) activity monitor is a combined uniaxial accelerometer and heart rate monitor, which was used to measure accelerometer-derived physical activity. The Actiheart is a reliable and valid measurement method, and was deemed appropriate for our target population, because the combination of accelerometer data with heart rate data would be better able to determine the intensity of physical activities [23]. The Actiheart was attached to the participation’s chest by using two Electrocardiography (ECG) electrodes. The Actiheart is a lightweight (8gram) and compact (7×33mm) device, connected to the two ECG electrode and capable of storing time-sequenced data. Acceleration (1D, vertical axis) was measured with a 15-second epoch by a piezoelectric element within the unit with a frequency range of 1-7Hz. The Actiheart output provides activity counts and heart rate data per minute, simultaneously.

### Activity energy expenditure (Actiheart)

Based on the Actiheart data, AEE estimates in kcals kg^−1^ min^−1^ were calculated for each minute by combining activity counts and heart rate in a branched equations model as described in literature [23,24] and as proposed by the Actiheart software for AEE (see supplemental file). A branched equation model allows the Actiheart to accurately assess AEE even when there is low body movement, but high heart rate during an activity. The combined activity and heart rate algorithm to calculate AEE needs the individual’s sleeping heart rate. The sleeping heart rate was calculated by averaging the minute-to-minute heart rate between 2.00-5.00am on the first day the Actiheart was worn.

When heart rate was missing, AEE was calculated based on the activity algorithm only for the specific missing minute. The total AEE per week was calculated by summing up the AEE minute-to-minute data, divided by the number of valid days the Actiheart was worn and multiplied by seven (assuming that the average amount of physical activity a day is representative for all weekdays and weekend days).

In addition, MET values were calculated for each minute based on the AEE minute-to-minute data, following the Ainsworth’ compendium of physical activities [20]. In the next step, MET values per minute were categorised in the following MET categories: sedentary behaviour(1.0–1.5 METs), light intensity (1.6-2.9 METs), moderate intensity (3.0-5.9 METs) and vigorous intensity (≥6 METs). Sum scores of all minutes in each MET category were calculated. Also, a sum score for al minutes of physical activity was calculated (≥1.6 METs). Sum scores were divided by the number of valid days the Actiheart was worn and multiplied by seven for week scores.

### Statistical analysis

Descriptive statistics were used to describe the demographic characteristics of the study population. Test-retest reliability of the Adapted-SQUASH was determined by calculating Intraclass correlation coefficients (ICCs) (two-way random, absolute agreement, single measures) for the total activity score (total, four main domains separately and all individual item separately), as well for the total minutes of activity (total and separately per intensity category) between the first and second measurement occasion. The ICC quantifies the degree to which the two measurements are absolutely related [25]. Since there is no widely accepted criterion for defining the strength of a correlation, we used a general guideline for clinical research: a correlation below 0.25 indicates little or no agreement, a correlation between 0.25 and 0.50 indicates fair agreement, a correlation between 0.50 and 0.75 indicates moderate to good agreement and a correlation higher than 0.75 indicates good to excellent agreement [26]. Also, confidence intervals were calculated for the ICCs. Additionally, Bland-Altman analyses were performed to illustrate the agreement between the first and second measurement of the Adapted-SQUASH [27,28]. Subsequently, a one-sample t-test was performed to determine any systematic bias.

Concurrent validity of the Adapted-SQUASH was determined by calculating a Spearman correlation coefficient between the total activity score (min x intensity/week) based on the baseline administration of the Adapted-SQUASH and the total AEE (kcals kg^−1^/week) based on the Actiheart data. Non-parametric Spearman correlation coefficients were chosen because assumptions of normality were not met for the outcomes of the Adapted-SQUASH and the two continuous outcome variables do not have the same measurement unit. In addition, concurrent validity of the Adapted-SQUASH was determined by calculating an ICC between the total minutes of activity (min/week) based on the baseline administration of the Adapted-SQUASH and the total minutes of activity (min/week) based on the Actiheart data, and by performing a Bland-Altman analysis. Although ICCs are preferred if the two measurement instruments are expressed in the same units (min/week) [29], a Spearman correlation coefficient was also calculated between the total minutes of activity assessed with the Adapted-SQUASH and Actiheart to compare our correlation with previous literature [4,11,13,14]. There is no consensus on how high correlations should be in order to demonstrate acceptable validity of a physical activity questionnaire [30]. The same interpretation of correlations as mentioned above is used for the validity of the Adapted-SQUASH. The level of significance was set at 0.05. Data were analysed using the Statistical Package for the Social Science (IBM SPSS Statistics, version

24).

## Results

A convenience sample of adults with disabilities (N = 80) was approached. Finally, 68 participants were included in the test-retest reliability study and 58 in the validity study. Twelve out of 80 participants were excluded from the test-retest reliability study because they did not fill out the second questionnaire due to illness (N = 1), surgery (N = 1) or unknown reasons (N = 10). Based on the characteristics, the included and excluded sample only statistically significantly differed in average body weight (see supplemental file). Body weight was on average 89.8±3.2 kg in the excluded sample and 79.1±14.7 kg in the included sample. Based on the criterion of a minimum of four days valid Actiheart accelerometer data, 22 out of 80 participants were excluded from the validity study. We included participants with four days (N = 5), five days (N = 1), six days (N = 6), and seven days (N = 46) of valid Actiheart accelerometer data, whereof all participants had at least three weekdays and one weekend day available. Based on the characteristics, the included and excluded sample only significantly differed in the use of mobility aid (see supplemental file). In the excluded sample, more people used a mobility aid (32%) compared to the included sample (17%).

The characteristics of the participants for the test-retest reliability (N = 68) and the validity (N = 58) studies are presented in Table 1. The Adapted-SQUASH was completed for a second time after a mean period of 17±4 days.

**Table 1.** Characteristics of the participants for the test-retest reliability study (N = 68) and the validity study (N = 58)

|  | **Test-retest reliability**<br>**study (N=68)** | **Validity study**<br>**(N=58)** |
| --- | --- | --- |
|  | Mean±SD or N(%) | Mean±SD or N(%) |
| Gender (% male) | 31 (46) | 27 (47) |
| Age (years) | 56.9 ± 17.6 | 54.7 ± 18.7 |
| Body height (m) | 1.73 ± 0.10 | 1.74 ± 0.09 |
| Body weight (kg) | 79.1 ± 14.7 | 80.8 ± 14.3 |
| Body Mass Index (kg/m<sup>2</sup>) | 26.6 ± 4.7 | 26.7 ± 4.5 |
| Drug use (% yes) | 60 (88) | 48 (83) |
| Use of mobility aid (% yes) | 17 (25) | 10 (17) |
| Diagnosis |  |  |
| Musculoskeletal disease | 2 (3) | 2 (3) |
| Brain disorder | 22 (32) | 15 (26) |
| Neurologic disease | 5 (7) | 5 (9) |
| Organ disease | 31 (46) | 29 (50) |
| Other diseases<sup>a</sup> | 6 (9) | 7 (12) |

SD=standard deviation, N=number of participants
<sup>a</sup> E.g. Ménière's disease, Hereditary Motor and Sensory Neuropathy type II, worn neck vertebrae, low-back pain.

### Test-retest reliability

The ICC for the repeated Adapted-SQUASH measurements was 0.67 (p< 0.001) for the total activity score, and 0.76 (p< 0.001) for the total minutes of activity per week, which respectively indicated a moderate to good and good to excellent agreement [25] (Table 2). Test-retest reliability within the light, moderate and vigorous intensity categories were respectively 0.89 (p< 0.001), 0.64 (p< 0.001), and 0.32 (p = 0.004). ICCs for the separate activity categories were: 0.39 (p< 0.001) for commuting activities, 0.77 (p< 0.001) for activities at work, 0.41 (p< 0.001) for household activities, and 0.44 (p< 0.001) for leisure-time activities. Test-retest reliability of the separate items of the questionnaire ranged from 0.00 for intense activities at work to 0.81 for walking during commuting (Table 2). Test-retest reliability of the new added items for handcycling activities during commuting and leisure time and wheelchair riding during commuting could not be determined because too few participants reported this activity. Test-retest reliability for wheelchair riding in leisure time was 0.27 (p = 0.011).

**Table 2.** Intraclass correlation coefficients (ICC) between the first and second measurement of the Adapted-SQUASH (N = 68)

|  | **Physical activity levels** |  | **Test-retest reliability** |  |  |
| --- | --- | --- | --- | --- | --- |
|  | First test | Second test | ICC | 95%CI | p |
|  | Median (IQR) | Median (IQR) |  |  |  |
| **Main outcomes** |  |  |  |  |  |
| Total activity score<sup>a</sup> | 1706 (658 – 4151) | 1950 (900 – 3864) | .67 | .51-.78 | <.001 |
| Total minutes of activity/week | 379 (189 – 861) | 473 (246 – 939) | .76 | .64-.85 | <.001 |
| **Intensity categories (min/week)** |  |  |  |  |  |
| Light | 86 (30 – 233) | 90 (30 – 278) | .89 | .83-.93 | <.001 |
| Moderate | 68 (3 – 308) | 119 (30 – 366) | .64 | .48-.76 | <.001 |
| Vigorous | 53 (30 – 131) | 78 (30 – 135) | .32 | .09-.51 | .004 |
| **Item activity scores<sup>a</sup>** |  |  |  |  |  |
| Commuting | 0 (0 – 10) | 0 (0 – 15) | .39 | .17-.57 | <.001 |
| Walking | 0 (0 – 0) | 0 (0 – 0) | .81 | .72-.88 | <.001 |
| Bicycling | 0 (0 – 8) | 0 (0 – 0) | .29 | .06-.50 | .007 |
| Handcycling | 0 (0 – 0) | 0 (0 – 0) |  | NA |  |
| Wheelchair riding | 0 (0 – 0) | 0 (0 – 0) |  | NA |  |
| Activities at work | 0 (0 – 705) | 0 (0 – 525) | .77 | .65-.85 | <.001 |
| Light | 0 (0 – 705) | 0 (0 – 450) | .78 | .67-.86 | <.001 |
| Vigorous | 0 (0 – 0) | 0 (0 – 0) | .00 | -.24-.24 | .499 |
| Household activities | 53 (21 – 116) | 54 (16 – 112) | .41 | .20-.59 | <.001 |
| Light | 45 (16 – 60) | 33 (15 – 75) | .43 | .21-.61 | <.001 |
| Vigorous | 0 (0 – 15) | 0 (0 – 30) | .20 | -.03-.42 | .044 |
| Leisure time | 136 (73 – 238) | 178 (91 – 244) | .44 | .23-.61 | <.001 |
| Walking | 19 (0 – 45) | 21 (8 – 45) | .21 | -.03-.42 | .046 |
| Bicycling | 15 (0 – 30) | 24 (0 – 38) | .15 | -.09-.38 | .110 |
| Handcycling | 0 (0 – 0) | 0 (0 – 0) |  | NA |  |
| Wheelchair riding | 0 (0 – 0) | 0 (0 – 0) | .27 | .04-.48 | .011 |
| Gardening | 0 (0 – 28) | 0 (0 – 30) | .19 | -.05-.41 | .059 |
| Odd jobs | 0 (0 – 6) | 0 (0 – 26) | .50 | .30-.66 | <.001 |
| Sports | 45 (0 – 105) | 45 (23 – 105) | .76 | .64-.85 | <.001 |

IQR=Inter Quartile Range, ICC=Intraclass Correlation Coefficient, CI=Confidence Interval, NA=not applicable due to too low response on this item.
<sup>a</sup> Activity score = minutes x intensity

Bland-Altman analyses showed that the mean difference between the first and second measurement was not significantly different from zero for both the total activity score (t_67_ = −0.03, p = 0.98) and for the total minutes of activity (t_67_ = 0.11, p = 0.92), indicating no systematic bias between the two measurements. We found wide Limits of Agreement (LOA) with 95% of the measurements of the total activity score within the boundaries of 4072 activity score above and below the mean difference (Fig 1), and with 95% of the measurements of the total minutes of activity within the boundaries of 945 minutes activity above and below the mean difference (Fig 2). Besides, based on the Bland-Altman plots the absolute amount of time spent on physical activity and the total activity score were higher at the second measurement occasion than at the first measurement occasion, while the total activity score was lower at the second measurement than at the first measurement occasion.

**Fig 1.**
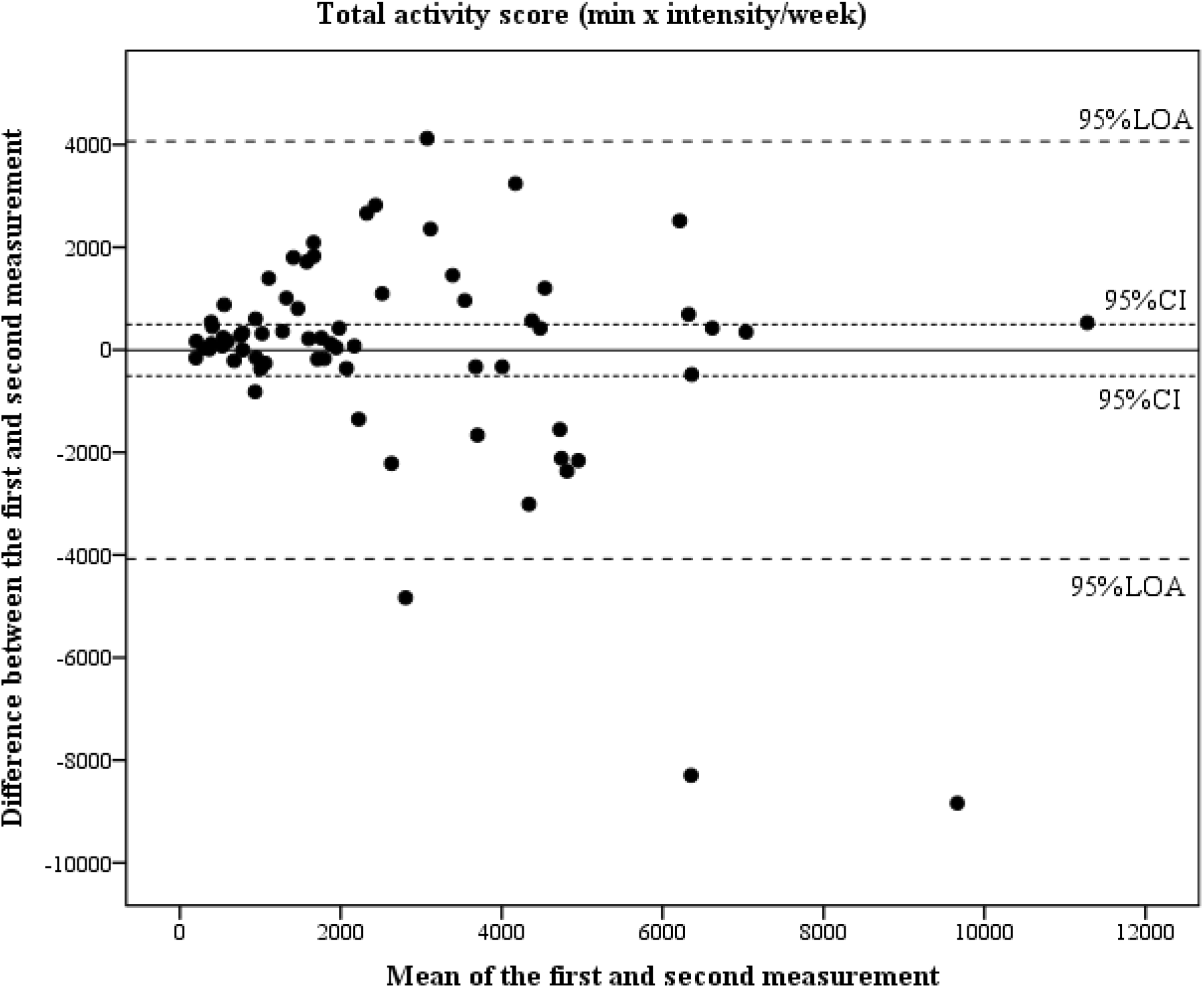
The differences between the total activity scores on the first and second measurement of the Adapted-SQUASH, plotted against their mean for each participant, together with the 95% confidence interval (CI) and the 95% Limits of Agreement (LOA) (N = 68).

**Fig 2.**
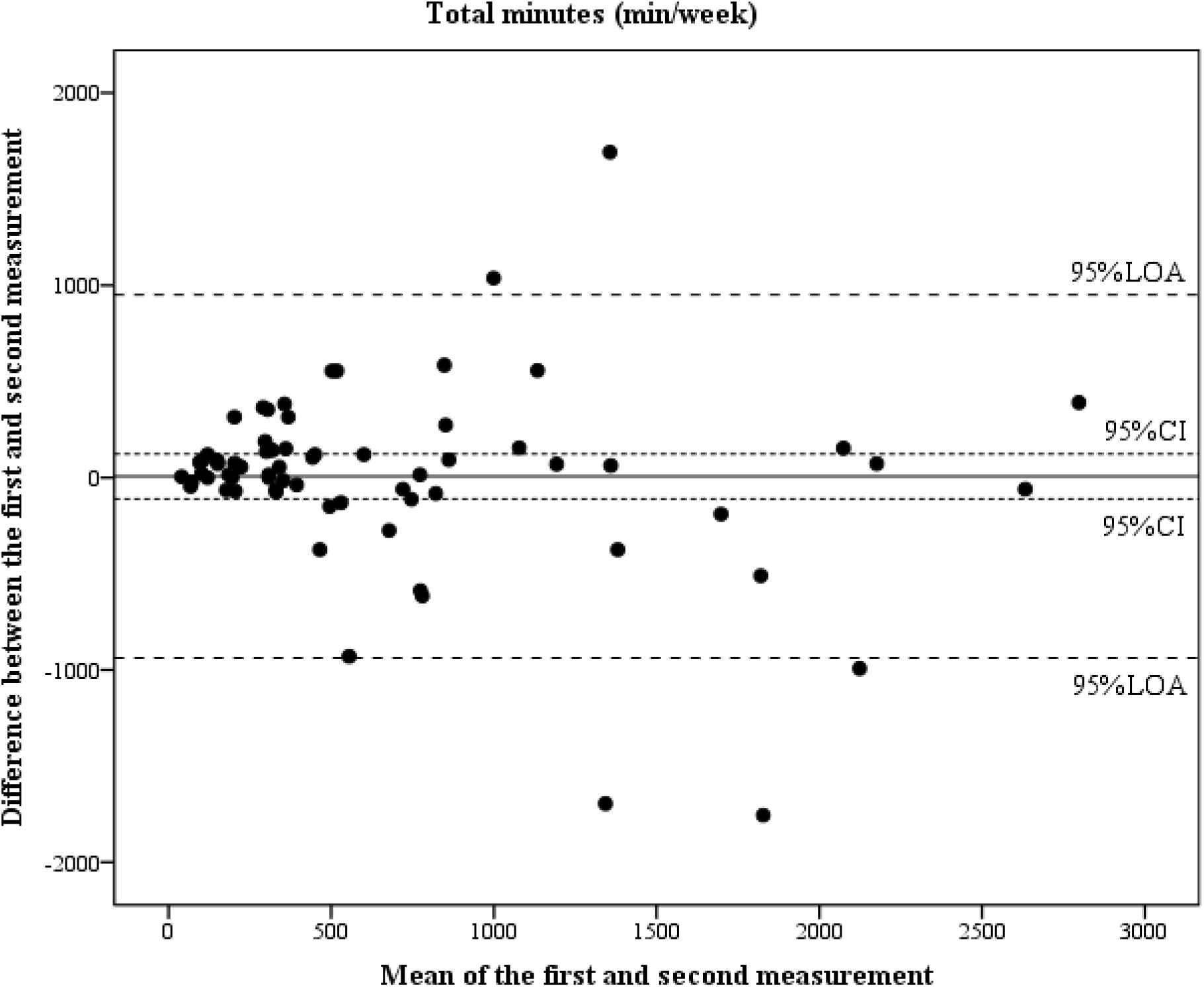
The differences between the total minutes of activity on the first and second measurement of the Adapted-SQUASH, plotted against their mean for each participant, together with the 95% confidence interval (CI) and the 95% Limits of Agreement (LOA) (N = 68).

### Concurrent validity

Correlation coefficients for the concurrent validity are presented in Table 3. A significant Spearman correlation coefficient was found between the total activity score from the Adapted-SQUASH and the AEE from the Actiheart (ρ = 0.40, p = 0.002). A significant ICC of 0.22 was found between the total minutes of activity per week from the Adapted-SQUASH and the total minutes of activity per week from the Actiheart (p = 0.027). The correlation coefficients indicated fair and little agreement, respectively. No significant ICCs were found between the total minutes of light and moderate activity per week calculated with the Adapted-SQUASH and Actiheart. Only a significant ICC of 0.21 (p = 0.046) was found between the total minutes of vigorous activity per week from the Adapted-SQUASH and Actiheart, indicating little agreement between the two measurement tools.

Bland-Altman analysis showed that the mean difference between the total minutes of activity calculated with the Adapted-SQUASH and Actiheart was significantly different from zero (t_57_ = 3.48, p = 0.001), indicating systematic bias between the two. We found wide LOA with 95% of the measurements of the total minutes of activity within the boundaries of 1485 minutes above and below the mean difference (Fig 3). Besides, based on the Bland-Altman plot the absolute amount of time spent on physical activity was higher reported in the Adapted-SQUASH questionnaire compared to physical activity assessed with the Actiheart.

**Table 3.**
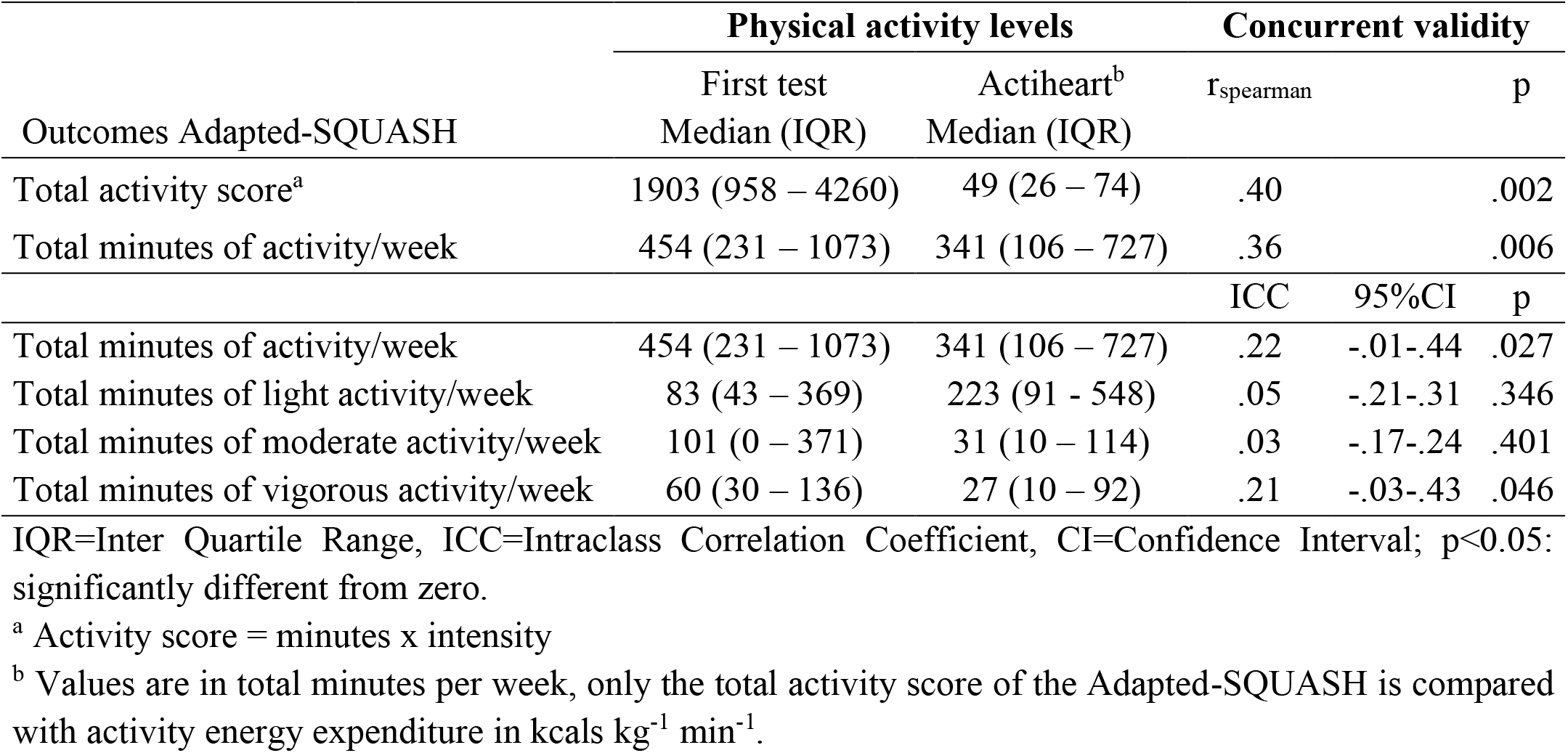
Correlation coefficients between the first measurement of the Adapted-SQUASH and the Actiheart (N = 58)

| Outcomes Adapted-SQUASH | Physical activity levels |  | Concurrent validity |  |  |
| --- | --- | --- | --- | --- | --- |
| | First test<br>Median (IQR) | Actiheart<sup>b</sup><br>Median (IQR) | $r_{\text{spearman}}$ | | p |
| Total activity score<sup>a</sup> | 1903 (958 – 4260) | 49 (26 – 74) | .40 |  | .002 |
| Total minutes of activity/week | 454 (231 – 1073) | 341 (106 – 727) | .36 |  | .006 |
|  |  |  | ICC | 95%CI | p |
| Total minutes of activity/week | 454 (231 – 1073) | 341 (106 – 727) | .22 | -.01-.44 | .027 |
| Total minutes of light activity/week | 83 (43 – 369) | 223 (91 - 548) | .05 | -.21-.31 | .346 |
| Total minutes of moderate activity/week | 101 (0 – 371) | 31 (10 – 114) | .03 | -.17-.24 | .401 |
| Total minutes of vigorous activity/week | 60 (30 – 136) | 27 (10 – 92) | .21 | -.03-.43 | .046 |

IQR=Inter Quartile Range, ICC=Intraclass Correlation Coefficient, CI=Confidence Interval; p<0.05: significantly different from zero.
<sup>a</sup> Activity score = minutes x intensity
<sup>b</sup> Values are in total minutes per week, only the total activity score of the Adapted-SQUASH is compared with activity energy expenditure in $\text{kcal kg}^{-1} \text{ min}^{-1}$ .

**Fig 3.**
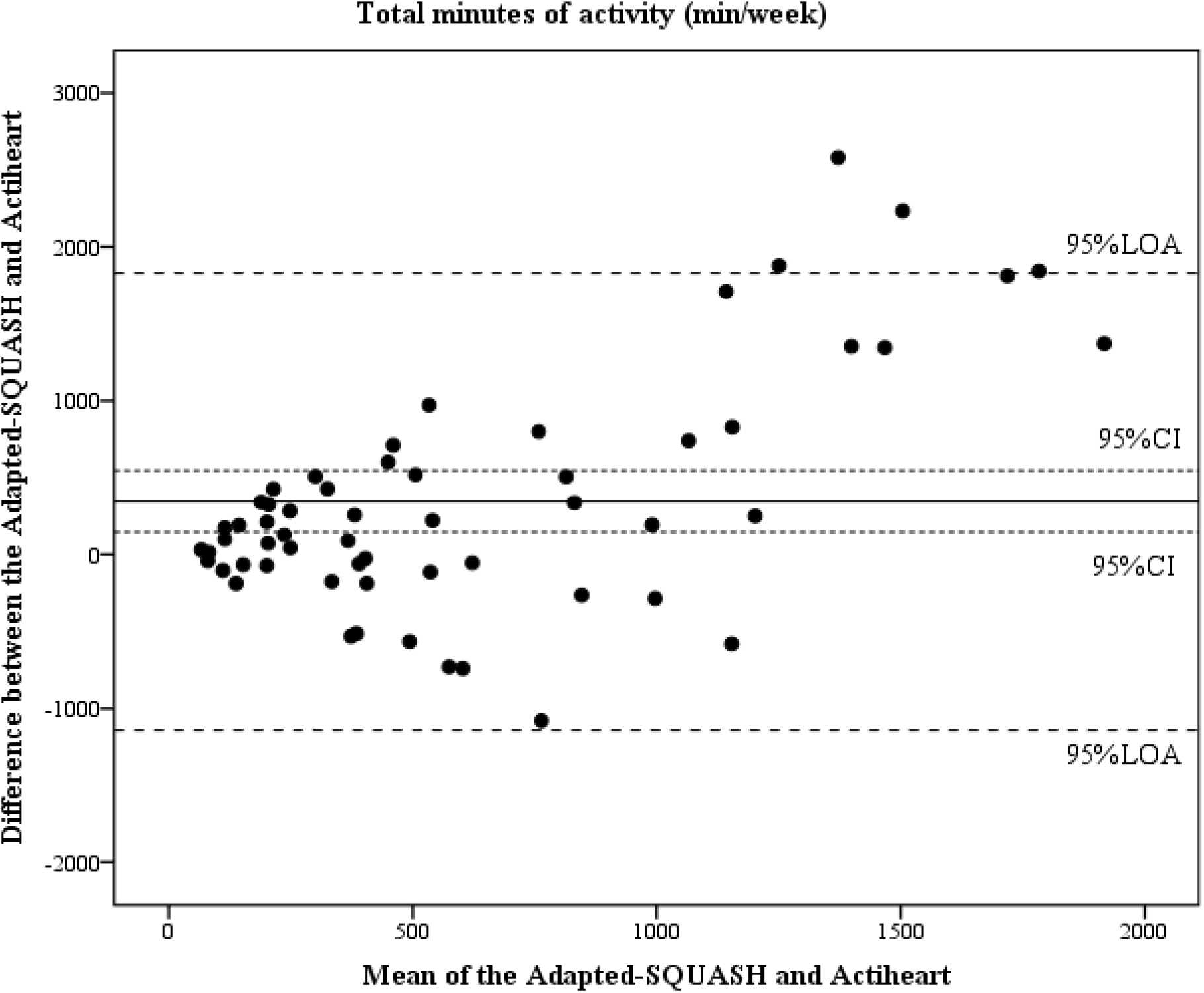
The differences between the total minutes of activity calculated with the Adapted-SQUASH and Actiheart, plotted against their mean for each participant, together with the 95% confidence interval (CI) and the 95% Limits of Agreement (LOA). (N = 58).

## Discussion

The current study showed good reproducibility of the Adapted SQUASH to assess self-reported physical activity in populations of people with disabilities, but not at the individual level since the Bland-Altman analyses found wide LOA. In addition, the current study showed fairly acceptable validity of the Adapted SQUASH, but the Bland-Altman analysis showed wide LOA, which indicates that self-reported physical activity individually assessed with the Adapted-SQUASH does not accurately represent individually accelerometer-derived physical activity assessed with the Actiheart in people with disabilities.

### Test-retest reliability

The test-retest reliability of the total activity score per week (ICC = 0.67, p<.001) of the Adapted-SQUASH is slightly higher compared to the Spearman correlation coefficients found in studies of the original SQUASH among 50 healthy adults (ρ = 0.58) [4], among 44 patients after a total hip arthroplasty (ρ = 0.57) [13], but slightly lower compared to a study among 52 patients with ankylosing spondylitis (ρ = 0.89) [14]. Also, our result of the test-retest reliability of the total minutes per week (ICC = 0.76, p<.001) of the Adapted-SQUASH is comparable to the Spearman correlation coefficient for the test-retest reliability of the PASIPD in similar populations with a disability (ρ = 0.77) [11]. A special note when comparing the test-retest reliability of our study to others is that we examined the test-retest reliability by using ICCs, while others used Spearman correlation coefficients [4,13,14]. ICCs give lower correlation coefficients compared to Spearman correlation coefficients, because an ICC is the absolute agreement between the first and second measurement, which does not correct for systematic differences. In accordance with previous studies [13,14], the Bland-Altman analysis showed no systematic bias on total activity scores between test and retest. Although the Adapted-SQUASH has good test-retest reliability and the mean differences between the first and second measurement are close to zero, relatively wide LOA are found for the total activity score and the total minutes of activity, which indicated that the degree of repeatability is insufficient at the individual level and/or that levels of physical activity fluctuate over time. Therefore, the Adapted-SQUASH can be used to assess self-reported physical activity behaviour in large (patient) populations, but is not acceptable to monitor individual physical activity levels. Also, it indicates that large changes in the outcomes of the Adapted-SQUASH should be found when interested in the course of self-reported physical activity over time (e.g. before and after an intervention or treatment).

The Adapted-SQUASH also calculated the total minutes of light, moderate and vigorous activity per week. In previous literature, Wendel-Vos et al. (2003) and Wagenmakers et al. (2008) found the highest Spearman correlation coefficient for the total minutes of vigorous activity per week, respectively 0.92[4] and 0.85[13], while we found the lowest correlation for the total minutes of vigorous activity per week (ICC = 0.32, p = .004). Explanation of this outcome is that vigorous intensity activities, such as weekly scheduled sports activities, are the easiest to recall for healthy adults [31], while intermittent light intensity activities (e.g. walking) are more difficult to recall [32,33]. However, adults with disabilities might experience activities as more intense, since activities often cost more energy compared to healthy adults [16,17] and may be more variable over the day due to fatigue and lack of appropriate pacing behaviour [34–37]. Therefore, temporal fluctuation in light intensity activities in healthy adults, may be similar to temporal fluctuation in moderate or vigorous intensity activities in our target population. Furthermore, our sample reported less minutes of vigorous intensity activities (so a lower between subjects’ variance) compared to light intensity activities, which might give a lower ICC.

The Adapted-SQUASH provides information of different settings of physical activity (commuting activities, activities at work/school, household activities and leisure time activities including different sports). We found low test-retest reliability for leisure-time activities, which might be explained by the non-regular frequency of this type of activities per week, due to barriers to physical activity such as the amount of leisure-time, tiredness, or bad weather conditions [38]. The quite low correlation for intense activities at work could be due to a small percentage of the population who can perform intense activities at work and the high variability in vigorous activities. The two newly added items ‘wheelchair riding’ and ‘handcycling’ in the Adapted-SQUASH had low response, because our study excluded people who were completely wheelchair dependent. However, our study population did mention adapted sports in the category ‘sports activities’ (e.g. wheelchair basketball).

Another interesting variable is the sport outcome measure indicating good test-retest reliability (ICC = 0.76, p< 0.001), probably because sports activities are often easy to recall, and sports participation is a stable behaviour with scheduled regular practice. This variable is often used in clinical settings, as well as in policy making and governmental guidelines worldwide. Insight in sports activities can be used for a tailored advice regarding an active lifestyle during or after rehabilitation, which has health-influencing effects, is crucial for quality of life, mobility and participation in everyday life and is strongly recommended for adults with disabilities [39].

### Concurrent validity

The concurrent validity of the total activity score per week of the Adapted-SQUASH (ρ = 0.40, p = .002), when compared with the total AEE per week assessed with the Actiheart, is lower compared to the Spearman correlation coefficients found in studies of the original SQUASH among 50 healthy adults (ρ = 0.45, physical activity was assessed with the computer science and applications activity monitor) [4], among 44 patients after a total hip arthroplasty (ρ = 0.67, physical activity was assessed with an Actigraph accelerometer) [13], but higher compared to a study among 52 patients with ankylosing spondylitis (ρ = 0.35, physical activity was assessed with an Actigraph accelerometer) [14]. Also, the concurrent validity of the total minutes of activity per week of the Adapted-SQUASH (ICC = 0.22, p = .027 and ρ = 0.36, p = .006), when compared with the total minutes of activity assessed with the Actiheart, is lower compared to the Spearman correlation coefficient found in the study of the original SQUASH among 50 healthy adults (ρ = 0.56) [4], but higher compared to the Spearman correlation coefficient for the validity of the PASIPD among people with disabilities (ρ = 0.30, physical activity was assessed with an Actigraph accelerometer) [11]. The lower concurrent validity of physical activity questionnaires in people with disabilities compared to healthy adults might be due to variation of the questionnaire and variation of the standard. Also, cognitive function, which is sometimes affected in people with disabilities, might influence the recall of activities and thereby might explain the differences between self-reported and accelerometer-derived physical activity [32].

In addition, although the Bland-Altman analysis showed no systematic bias between the total minutes of activity per week assessed with the Adapted-SQUASH and Actiheart, the LOA were wide. This indicated that the Adapted-SQUASH does not accurately represent accelerometer-derived physical activity assessed with the Actiheart in individuals with disabilities. Previous literature also found that individual self-reported physical activity compared to physical activity assessed with an accelerometer was not accurate in people after joint arthroplasty [40] and in people with spinal cord injury [7]. Besides, the mean difference between the Adapted-SQUASH and Actiheart was 346 minutes per week, which indicates that people with disabilities seem to overestimate their self-reported physical activity assessed with the Adapted-SQUASH compared to accelerometer-derived physical activity assessed with the Actiheart. This is in agreement with previous literature [41–43]. This overestimation of actual time spent being physically active is probably attributable to recall bias, such as the difficulty in recalling short breaks during physical activity (e.g. socializing or refreshment during the reported time doing sports, or taking rest during the reported time doing gardening or household activities) [7], while the Actiheart does measure all sorts of short breaks during physical activity and over the day. Another potential bias between self-reported and accelerometer-derived physical activity outcomes may reside in the appreciation and perception of physical activities and their intensities, which notions may be quite different in our population in the context of their often low physical work capacity [44] and phenomena of fatigue during the day [35,36]. This introduces a difference in what one does and what one perceives.

Consequently, for the total minutes of vigorous activities per week low or little agreement was found between the Adapted-SQUASH and Actiheart (ICC = 0.21, p = .046), while no agreement was found for the total minutes of light (ICC = 0.05, p = .346) and moderate (ICC = 0.03, p = .401) activities per week. This suggests that the perceived intensity of activities in people with disabilities is not in agreement with the accelerometer-derived intensities of activities assessed with the Actiheart. Therefore, we suggest to use the total minutes of physical activity per week assessed with the Adapted-SQUASH when interested in dose-response relationships among for instance physical activity and health outcomes, or between physical activity and the received intervention/treatment in people with disabilities.

### Limitations

A few limitations need to be considered. First, the Adapted-SQUASH used MET values from the Ainsworth compendium of physical activities, which were derived from and intended for use in able-bodied adults [20]. This limitation could have overestimated the total activity score for each intensity category [20,45], because our target population probably experiences activities as more intense compared to healthy adults [16,17], as well as less consistent during the day. Also, the Adapted-SQUASH is sensitive to overestimation of frequency and/or duration of the activities, due to recall bias. A more or less similar limitation is however true for the Actiheart device, where the used sensor algorithms are not specific to people with disabilities, but have been derived from the general healthy population [23,24]. This stresses the need for more population-specific validation studies also of objective physical activity measurement tools in the future [46].

Thirdly, the test-retest period was on average seventeen days. This duration could be too short to prevent participants from copying the Adapted-SQUASH from memory. However, following the recommendations of Matthews et al. (2012), we have consciously chosen for this short recall period to decrease the reporting error of activities, since physical activity levels tend to fluctuate between days and weeks due to weather conditions [47] and/or due to fluctuating experienced health or fatigue conditions among this population of persons with a disability [37]. Furthermore, we did not check at the participant if the week the Actiheart was worn was a representative week of their physical activity behaviour.

Lastly, the Actiheart is a device capable of measuring heart rate and acceleration, and combines these variables in a branched equation model to calculate AEE [23,24]. However, we found a large amount of missing heart rate data in our sample, while calculating AEE based on the heart rate and combined algorithm is preferred [24]. The median percentage of missing heart rate was 22% (inter quartile range: 10% - 42%). The unsuccessful measurement of heart rate may have happened due to malfunction of the battery or the electrodes. However, if during the week participants felt that the electrodes loosened or if the electrodes had not been replaced by the fourth day, the instruction was given to replace the electrodes. As stated above another limitation is that the algorithm from the Actiheart to calculate AEE has not been validated among adults with deviating movement patterns and adults using drugs against high blood pressure, who are included in our target population. This is however the case for most of the activity monitor devices currently available [48].

### Practical implications and further research

The Adapted-SQUASH provides information on various dimensions (frequency, duration and intensity) and settings (e.g. household, leisure time), is inexpensive, and has low burden for participants to fill in. Together this turns the Adapted-SQUASH into a useful tool to assess self-reported physical activity among adults with disabilities in large population studies. Firstly, the Adapted-SQUASH can be used in community and health-care settings, like rehabilitation centres, to monitor physical activity levels in large heterogeneous populations with disabilities. For this practical use, the Adapted-SQUASH is distinctive compared with other physical activity questionnaires (e.g. PASIPD), because even though the questionnaire specifically assesses type, frequency and intensity of activities, it is short and quick to fill in and it includes physical activities for wheelchair users and adapted sports. Secondly, the Adapted-SQUASH can be used for large longitudinal cohort studies or intervention studies to evaluate self-reported physical activity. For example, the Adapted-SQUASH has already been used in the longitudinal cohort study ReSpAct, which aimed to evaluate physical activity in people with disabilities during and after a physical activity stimulation programme [8,9]. When accurate and complete measures of physical activity are preferred in further research among large populations with disabilities, we suggest using both the Adapted-SQUASH (in the total sample) and an activity monitor (in a sub-sample). The Adapted-SQUASH provides information on the setting of the activity, while an activity monitor provides information on intermittent activities (e.g. walking at home and taking rest during activities) [7,33]. So, selection of the best measurement to assess physical activity depends on the purpose, construct, measurement unit, population, setting etc.[6].

For practical implications, we recommend using the total minutes of activity per week or the total activity score, which were the two main outcome measures of the Adapted-SQUASH, to assess self-reported physical activity in people with disabilities. The test-retest reliability of the total minutes of activity per week was good but systematic bias with the Actiheart was found. The test-retest reliability of the total activity score per week was lower and the perceived intensity of activities (light, moderate and vigorous) was not in agreement with the Actiheart. However, outcomes should be interpreted with caution since our sample of people with disabilities overestimated their physical activity. Also, for future research it is recommended to assess the validity and test-retest reliability of the Adapted-SQUASH among people who are completely wheelchair dependent.

## Conclusion

The Adapted-SQUASH is an acceptable measure to assess self-reported physical activity in large populations of people with disabilities but is not applicable at the individual level due to the wide LOA. Self-reported physical activity assessed with the Adapted-SQUASH does not accurately represent accelerometer-derived physical activity assessed with the Actiheart in individuals with disabilities. They seem to overestimate their physical activity and find it difficult to recall the perceived intensity of the activity. The test-retest reliability and concurrent validity of the Adapted-SQUASH are comparable to other physical activity questionnaires among people with disabilities. We recommend using the total minutes of activity per week and/or total activity score, derived from the Adapted-SQUASH, to evaluate physical activity in large populations of people with disabilities in rehabilitation practice and (cohort) research.

## Data Availability

The data referred to in the manuscript is not yet available.

## Acknowledgements

The authors wish to thank the participating subjects and the students in the bachelor and master Human Movement Sciences at the University of Groningen who contributed to the data collection. The authors would like to thank Leonie A. Krops for her critical reading and comments on a draft of the manuscript.

